# Cohort profile: the SEROCoV-KIDS population-based cohort study and biobank on the impact of the COVID-19 pandemic in children and adolescents in Geneva, Switzerland

**DOI:** 10.64898/2026.01.14.26344108

**Authors:** Elsa Lorthe, Andrea Loizeau, Viviane Richard, Roxane Dumont, María-Eugenia Zaballa, Francesco Pennacchio, Julien Lamour, Arnaud G L’Huillier, Hélène Baysson, Natalia Fernandez Clares, Nicolas Bovio, Mayssam Nehme, Pierre Lescuyer, Nicolas Vuilleumier, Klara M. Posfay-Barbe, Rémy P. Barbe, the SEROCoV-KIDS study group, Idris Guessous, Silvia Stringhini

**Affiliations:** Division of Primary Care Medicine, Geneva University Hospitals, Geneva, Switzerland; Université Paris Cité, Inserm, Inrae, Obstetric, Perinatal, Paediatric Life Course Epidemiology (OPPaLE) Research Team, Center for Research in Epidemiology and Statistics (CRESS), Paris, France; Geneva School of Health Sciences, HES-SO University of Applied Sciences and Arts Western Switzerland, Geneva, Switzerland; Pediatric Infectious Diseases Unit, Department of Women, Child and Adolescent Medicine, Geneva University Hospitals and Faculty of Medicine, Geneva, Switzerland; Laboratory of Virology, Department of Diagnostics, Geneva University Hospitals; Geneva, Switzerland; Department of Pediatrics, Gynecology and Obstetrics, Faculty of Medicine, Geneva, Switzerland; Department of Health and Community Medicine, Faculty of Medicine, University of Geneva, Geneva, Switzerland; Division of Laboratory Medicine, University Hospitals of Geneva, Geneva, Switzerland; Department of Medicine, Medical Faculty, Geneva University, Geneva, Switzerland; Division of General Pediatrics, Department of Woman, Child, and Adolescent Medicine, Geneva University Hospitals, Geneva, Switzerland; Division of Child and Adolescent Psychiatry, Department of Woman, Child, and Adolescent Medicine, Geneva University Hospitals, Geneva, Switzerland; School of Population and Public Health and Edwin S.H. Leong Centre for Healthy Aging, Faculty of Medicine, University of British Columbia, Vancouver, Canada

**Keywords:** COVID-19, cohort studies, child, adolescent, health, socioeconomic factors, health behavior

## Abstract

**Purpose:** The COVID-19 pandemic has had profound and multifaceted impacts on children and adolescents, exposing and deepening pre-existing inequalities while creating unique health, social, and educational challenges. In response to the limited evidence-based knowledge available, the SEROCoV-KIDS study was launched in 2021 as a prospective cohort and biobank to assess the pandemic’s effects on youth health and well-being. It focuses on the general population of Geneva, Switzerland, as well as subgroups with vulnerabilities, including prior SARS-CoV-2 infection, chronic health conditions, and socioeconomic disadvantage.

**Participants:** A total of 2199 children and adolescents, aged 6 months to 17 years, from 1340 households were enrolled in the SEROCoV-KIDS cohort, 2048 from the general population and 151 from clinics (i.e., children with chronic health conditions). At baseline, between December 2021 and June 2022, participants provided blood samples for serological testing and for long-term storage in the biobank, while comprehensive questionnaires were completed by a referent adult and adolescents aged 14 and older. Five follow-up online assessments were conducted until October 2025, addressing physical and mental health, development, quality of life, health behaviors, family dynamics, and education-related topics. The quantitative findings of the cohort study were enriched by a cross-sectional qualitative study conducted between October 2023 and March 2024, focusing specifically on socioeconomically disadvantaged populations.

**Findings to date:** In the population-based sample, 66.3% of participants tested seropositive for anti-SARS-CoV-2 nucleocapsid antibodies at baseline, and 4.1% reported symptoms consistent with post-COVID condition. Most children were minimally or not at all affected by the pandemic, showing good mental health over time. However, 8% of participants reported a positive pandemic impact, whereas 8–13% experienced negative impacts, mainly due to disrupted routines and reduced social support. Health behaviors like physical activity and sleep remained largely stable over the study period. Higher screen time at baseline was associated with poorer well-being. Children with chronic health conditions or experiencing socioeconomic and family disadvantage were disproportionately affected in terms of the health and psychosocial consequences of the pandemic.

**Future plans:** SEROCoV-KIDS demonstrates the value of child-focused cohorts for understanding the consequences of major societal events and for guiding evidence-based policy. Our next priority is to secure funding to prolong follow-up of this cohort and to maintain systematic surveillance of children and adolescents, so that emerging findings can directly inform public health and education policy over the coming years.

**Strengths and limitations of this study:**

- This large population-based pediatric cohort provides insights into the health and well-being of children and adolescents aged 6 months to 17 years at baseline, during and after the COVID-19 pandemic (from December 2021 to October 2025).
- This study integrates multifaceted findings on the general pediatric population and on subgroups of children with clinical and/or social vulnerabilities, combining quantitative and qualitative data to provide a deeper understanding of how the pandemic has affected their lives.
- This unique pediatric cohort and its associated biobank offer a rare opportunity to advance future pediatric research in Switzerland and abroad.
- Generalizability may be limited, as participating families tended to be more highly educated and somewhat more socioeconomically advantaged than the general Geneva population, despite nearly one-fifth reporting financial difficulties.
- The study was designed in response to the pandemic, and individual-level pre-pandemic data are lacking, which limits direct comparisons over time, relying instead on parent-reported perception of changes and impacts.

## Introduction

The COVID-19 pandemic has had far-reaching and multifaceted effects on global populations, with children and adolescents experiencing unique and evolving challenges across health, social, and educational domains. During the early stages of the pandemic, marked by high mortality rates and a sense of emergency, research primarily focused on understanding the pathophysiology of the disease in its acute phase, its transmission, diagnosis, treatment, and prevention. Preliminary evidence showed that children and adolescents had lower susceptibility to SARS-CoV-2 infection compared with adults, and played a lesser role than adults in transmission of SARS-CoV-2 at a population level.^1^ Little attention was given to the pediatric population, as children represented only a small fraction of diagnosed SARS-CoV-2 infections,^2,3^ and an even smaller fraction of severe cases.^3–5^ This was potentially linked to changes in contact patterns related to school closures, and underdiagnosis due to the scarcity of tests available, as well as the high frequency of asymptomatic infections or non-specific mild symptoms in children.^1^ However, with the emergence of variants of concern, it became evident that children could be infected and reinfected, and could transmit the infection to their households and classmates.^6–13^ Severe complications, such as multisystem inflammatory syndrome and post-COVID condition, were also increasingly reported, with a disproportionately higher burden observed among children with pre-existing health conditions.^3,14–16^

Beyond being a health crisis, the COVID-19 pandemic quickly became an economic and social crisis. Children and adolescents were largely affected, as their lives were profoundly disrupted by the extensive measures implemented to control the spread of the pandemic. School closures, social isolation, changes in family dynamics, unhealthy behaviors such as prolonged screen time or reduced physical activity, and generalized uncertainty posed significant challenges.^17–20^ This was particularly salient for older children and adolescents, for whom peer interactions are critical to development.^17,21^ Younger children were also highly vulnerable, as they faced prolonged stress during key developmental stages and often lacked the internal and external resources to cope adequately.^22–24^ While early meta-analyses showed only minimal increases in mental health symptoms among youth,^25,26^ subsequent studies reported a growing frequency of mental health issues, especially among girls and older adolescents, likely due to repeated lockdowns and lifestyle disruptions.^22,27–32^ However, not all outcomes were negative. In some cases, lockdowns fostered stronger family bonds and offered opportunities for more individualized learning experiences.^33,34^ Given these complex and evolving dynamics, it was essential to examine the pandemic’s mid- and long-term impact on the mental health and well-being of children and adolescents.

Since the earliest months of the pandemic, it became apparent that the pandemic not only revealed but also intensified existing social inequalities, disproportionately impacting disadvantaged populations.^35–37^ Adults from low-income, immigrant, or minority backgrounds were more likely to experience both direct (e.g., SARS-CoV-2 infection, severe COVID-19 disease, hospitalization, and death), and indirect (e.g., stress, mental health issues, job insecurity, financial difficulties) consequences of the pandemic.^35,38^ Among children and adolescents, factors such as poverty, low parental education, immigrant background, or racial/ethnic minority background were identified as risk factors for poor mental health outcomes,^39^ both prior to and throughout the COVID-19 pandemic.^17,40–42^ Gaining a deeper understanding of the social patterning of the pandemic long-term effects was essential for mitigating its enduring impact on youth well-being.

In addition to its psychological and social toll, the pandemic disrupted education, with school closures and remote learning contributing to worsening academic performance^43,44^ and widening educational disparities.^45^ While some children, particularly those from higher socioeconomic backgrounds, were able to maintain or even advance their learning through parental support and digital access, many disadvantaged students faced barriers such as limited internet access, lack of learning space, reduced educational support at home, and diminished access to school-based services.^46^ These disparities risk deepening pre-existing learning gaps and may have long-term consequences on academic achievement and future opportunities.

Recognizing the scarcity of data available at the beginning of the pandemic and anticipating the need for robust evidence to guide public health policies, in 2021 we launched SEROCoV-KIDS as a prospective cohort study and biobank to evaluate the impact of the COVID-19 pandemic on the health and well-being of children and adolescents. Its specific objectives were: (1) to assess the prevalence of anti-SARS-CoV-2 N and S antibodies in a pediatric population-based study; (2) to investigate the pandemic’s health and psychosocial effects on children and adolescents from the general population; and (3) to examine outcomes among subgroups with vulnerabilities, including prior SARS-CoV-2 infection, chronic health conditions, and/or socioeconomic disadvantage. We hypothesized that children’s health and well-being were both directly and indirectly affected by the pandemic, with disproportionately greater impacts observed in subgroups with clinical and/or social vulnerabilities.

## Cohort Description

### Study setting and context of the COVID-19 pandemic

The first confirmed COVID-19 case was reported in the canton of Geneva, Switzerland, on February 26^th^ 2020.^47^ In response to the surge in cases, strict measures were introduced on March 16^th^ 2020, including a partial lockdown and school closures until May 11^th^ 2020, followed by a gradual easing of the measures as cases declined. In later stages, despite experiencing multiple COVID-19 waves, Switzerland avoided school closures and prolonged or nationwide lockdowns, instead focusing on adult vaccination, COVID-19 certificates, mask mandates and remote work, with restrictions adjusted based on the epidemiological situation. Authorities prioritized keeping schools and certain activities open for children and adolescents whenever possible. By February 2022, most restrictions had been lifted, and all remaining measures were removed by May 2022. Switzerland launched large-scale vaccination campaigns in late December 2020, initially prioritizing high-risk groups such as the elderly, individuals with chronic conditions, and healthcare workers. The rollout gradually expanded to the broader population, with vaccination becoming available to children aged 12 and older in June 2021 and to those aged 5 and older in January 2022.^48^

### Cohort study design, population and recruitment procedures

This prospective cohort study of children and adolescents includes both a general population sample and a clinical sample of children with chronic health conditions (Figure 1). For the population-based sample, eligible children and adolescents were those living in the canton of Geneva and aged 6 months to 17 years at baseline. Participants were randomly selected from state registries, either specifically for this study or for other COVID-19 population-based seroprevalence studies conducted by our group.^47–50^ The registries were provided by the Swiss Federal Office of Statistics or the Cantonal Office for Population and Migration. Their siblings who met the eligibility criteria were also invited to participate. Children with chronic health conditions were referred by specialist physicians working at the Children’s Hospital (Geneva University Hospitals) in one of the following units: pneumology, immunology, allergology, gastroenterology, neurology, and rheumatology. They were eligible if they met the same age and residency criteria and had one or more medically diagnosed conditions lasting for at least six months. These included, but were not limited to, chronic respiratory or cardiac diseases, severe allergies, neurological disorders, rheumatic diseases, diabetes, inflammatory bowel disease, or immune system disorders.

**Figure 1.**
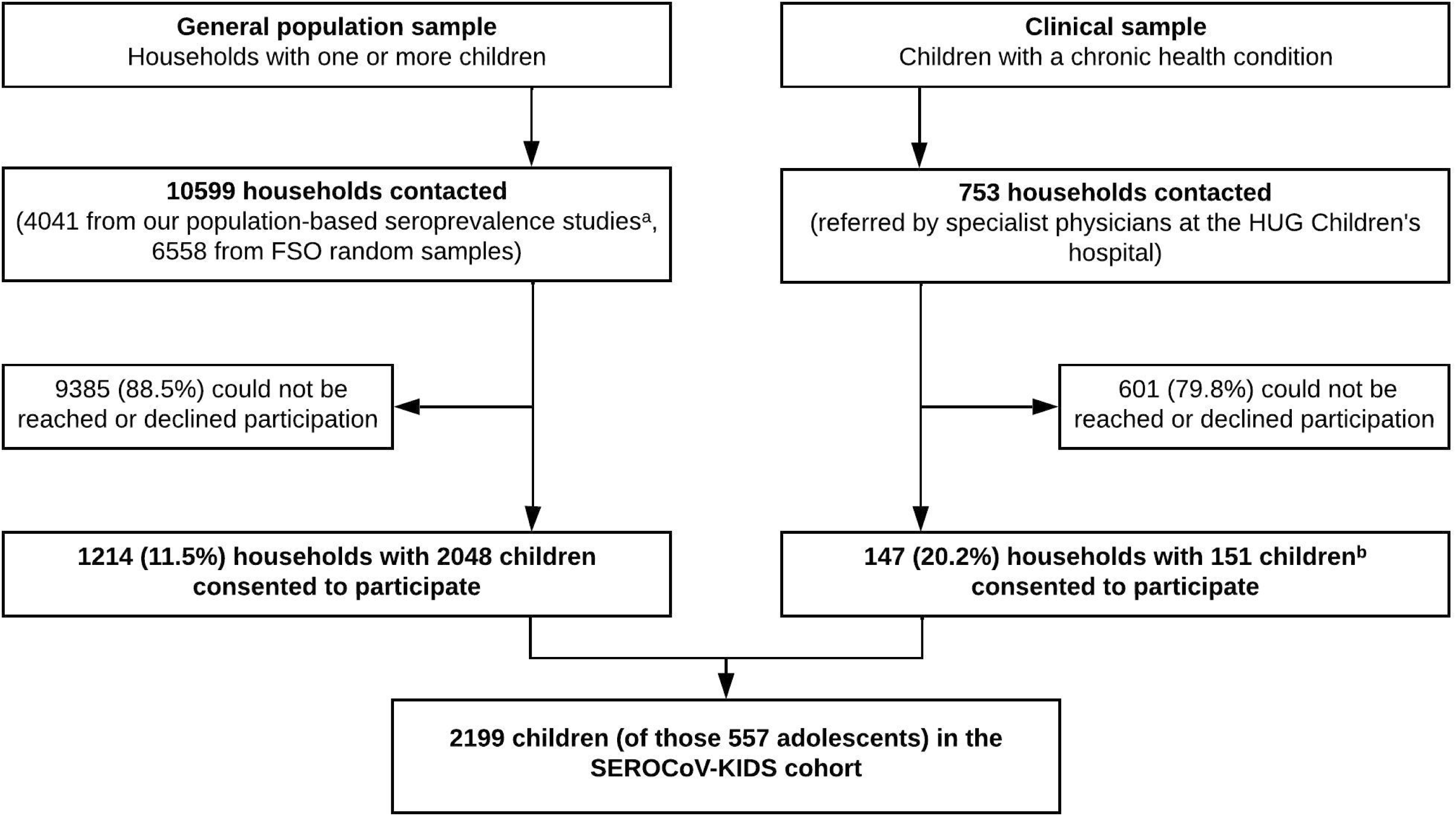
Flow chart of the study. Legend: Abbreviation: HUG, University Hospitals of Geneva. ^a^For 1802 former participants, the presence or absence of children in the household was unknown at the time of invitation. ^b^Proportions of children enrolled per hospital unit: 10 (6.6%) immunology, 63 (41.7%) pneumonolgy, 31 (20.5%) allergology, 10 (6.6%) gastro-enterology, 26 (17.2%) neurology, and 11 (7.3%) rheumatology.

A pilot phase was conducted in November 2021 among 130 children and adolescents of participants in a previous seroprevalence study,^51^ allowing us to validate all study procedures. Participants were enrolled in the general population sample between December 2021 and June 2022, and in the clinical sample between June and December 2022.

The Geneva Cantonal Commission for Research Ethics approved this study (N° 2021-01973). Adolescents from 14 years of age and parents (or legal guardians) of participants provided electronic or written informed consent at baseline. Children gave oral assent to participate. Participants who reached 14 years of age were individually invited to provide consent in June 2022 and in September 2023. Among them, 30 (49.2% of those contacted) agreed to complete their own questionnaires. Without opt-out requests, all participants were followed throughout the data collection period, including those transitioning into adulthood.

### Study procedures, blood collection and questionnaires

The baseline assessment included the completion of online inclusion questionnaires, and an optional serological test offered to all children and adolescents. To detect anti-SARS-CoV-2 antibodies, we used two highly accurate immunoassays: the Roche Elecsys anti-SARS-CoV-2 spike (S) and anti-SARS-CoV-2 nucleocapsid (N) assays (Roche Diagnostics, Rotkreuz, Switzerland). Seropositivity was determined using the manufacturer’s cut-off values: ≥0.8 U/mL for the S assay and index ≥1.0 for the N assay. COVID-19 exposure data were collected through a brief paper survey administered during the baseline visit.

An additional blood sample was collected from each child for long-term storage in the SEROCoV-KIDS biobank. All samples were labelled with unique identification numbers and stored at the Diagnostics Department of the Geneva University Hospitals. For participants whose referent adult provided consent for extended use of biospecimens, samples were preserved in the biobank for future analyses. Biobank regulation follows the guidelines of the Swiss Biobanking Platform.

At baseline, one referent adult per family completed a detailed digital questionnaire covering socio-demographic, health, lifestyle, well-being, and COVID-19 pandemic-related information for each participating child, as well as for themselves and their household. Adolescents aged 14 and older independently completed an age-appropriate questionnaire about their health and well-being. All questionnaires were completed online via the Specchio-Hub digital platform (https://www.specchio-hub.ch/),^52^ with a small number of participants opting for a paper-based format.

Annual follow-up questionnaires were administered in late spring 2023, 2024, and 2025 to monitor trends in physical and mental health, quality of life, development, and lifestyle over time. Validated scales were used whenever possible (Table 1, Table 2), and translated into French through a forward-backward translation approach if necessary. Additionally, thematic questionnaires were distributed in fall 2022 and spring 2024 to explore family- and education-related topics, respectively. At each time point, adolescents completed an age-appropriate questionnaire. All questionnaires were designed to be completed online within 15 to 30 minutes. In the case of non-response, up to three email reminders were sent, spaced 7 to 14 days apart, followed by a phone call and/or a paper reminder if needed. To encourage participation, families were entered into a raffle for small incentives (e.g., vouchers, movie tickets) after each annual questionnaire. Retention strategies also included regularly sharing study findings via email newsletters or webinars and distributing paper trifold flyers tailored to the young population.

**Table 1.**
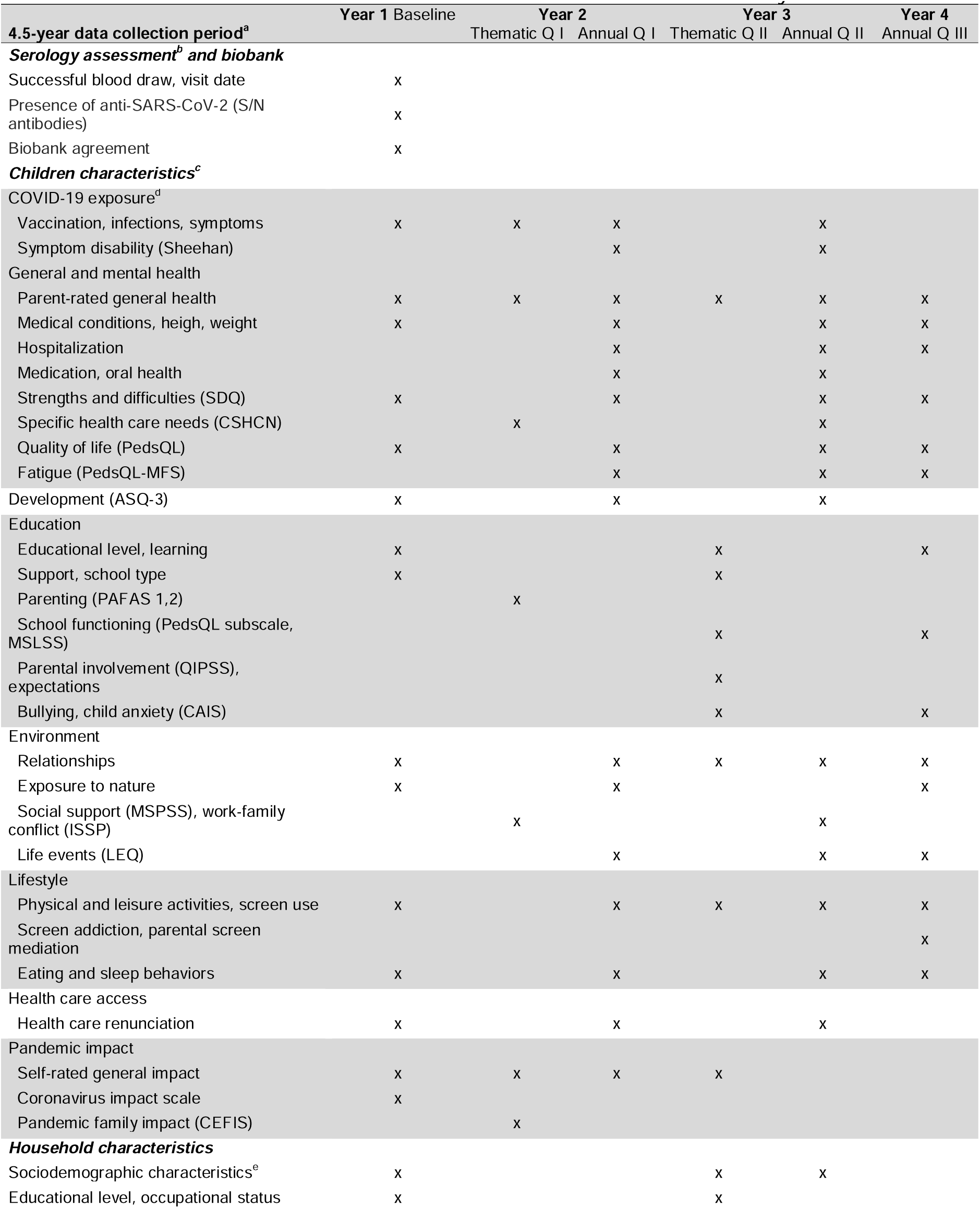
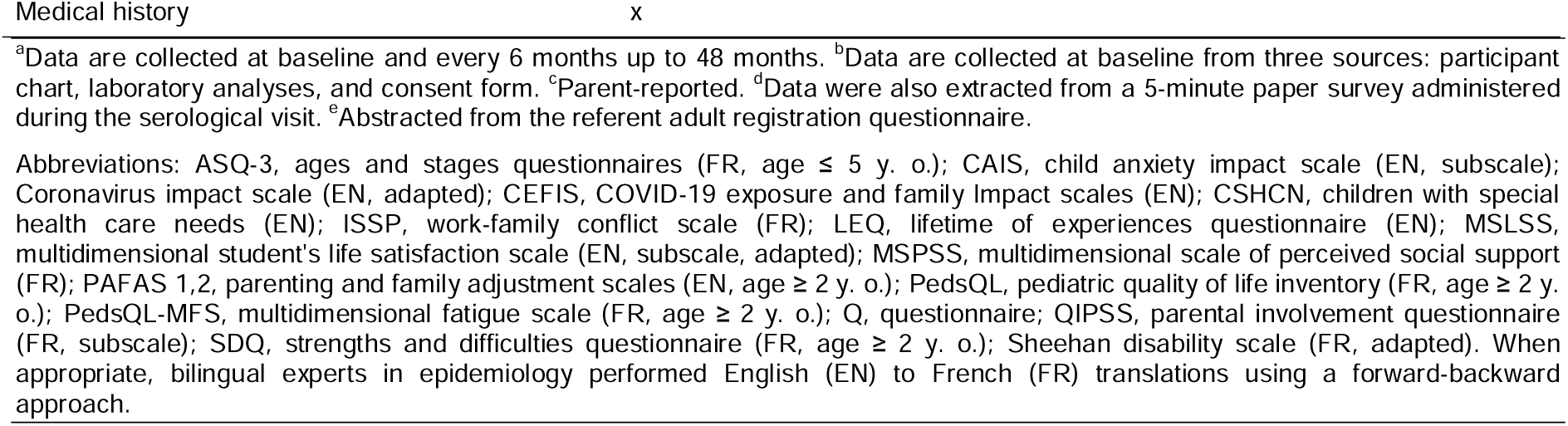
Children data collection over the course of the SEROCoV-KIDS study.

**Table 2.** Adolescents data collection over the course of the SEROCoV-KIDS study.

| 4.5-year data collection period<sup>a</sup> | Year 1 | Year 2 |  | Year 3 |  | Year 4 |
| --- | --- | --- | --- | --- | --- | --- |
|  | Baseline | Thematic Q I | Annual Q I | Thematic Q II | Annual Q II | Annual Q III |
| **Adolescent characteristics<sup>b</sup>** |  |  |  |  |  |  |
| COVID-19 exposure |  |  |  |  |  |  |
| Vaccination, perception | x |  |  |  |  |  |
| General and mental health |  |  |  |  |  |  |
| Self-rated general health | x |  |  | x | x | x |
| Height, weight |  | x |  |  | x | x |
| Gender identity, menstrual cycle |  |  |  |  | x |  |
| Quality of life (PedsQL), well-being (WHO-5) | x |  | x |  | x | x |
| Self-esteem (RSES) | x |  | x |  | x | x |
| Distress (IDPSQ-14), suicidal ideation | x |  | x |  | x | x |
| Depression (PHQ-9) |  |  |  |  | x | x |
| Self-perception |  | x | x |  | x |  |
| Personality (Big5) |  | x |  |  |  |  |
| Happiness (SHS), future prospects (FTP) |  | x |  |  |  |  |
| Acne (GEA, AcneQ4) |  |  | x |  |  |  |
| Resilience (BRS) |  |  | x |  | x | x |
| Education |  |  |  |  |  |  |
| Educational level, learning, support | x |  |  | x |  | x |
| School well-being (BASWBSS), burn-out (SBI) |  |  |  | x |  | x |
| Bullying (BVQr) | x |  | x | x |  |  |
| Environment |  |  |  |  |  |  |
| (Love) relationships | x | x | x |  | x | x |
| Social support (MSPSS) | x |  | x |  | x | x |
| Life events (PANALE) |  |  |  | x |  |  |
| Geopolitical awareness, eco-anxiety (HEAS) |  |  |  | x |  |  |
| Lifestyle |  |  |  |  |  |  |
| Physical activities (IPAQ) | x | x |  | x |  | x |
| Leisure activities, screen use | x |  | x | x | x | x |
| Artificial intelligence use |  |  | x | x |  | x |
| Gaming (IGCS) |  |  |  |  |  | x |
| Eating behaviors | x | x |  |  |  | x |
| Sleep behaviors (year 3, ISI-3) | x |  |  |  | x | x |
| Cyber-bullying, social networks (BSMAS) | x |  | x |  | x | x |
| Addictions | x |  | x |  | x | x |
| Pandemic impact |  |  |  |  |  |  |
| Self-rated general impact | x |  |  | x |  |  |

<sup>a</sup>Data are collected at baseline and every 6 months up to 48 months. <sup>b</sup>Self-reported. Abbreviations: AcneQ4, acne four-question scale (FR); Big5, big five-factor personality model (FR); BASWBSS, brief adolescents' subjective well-being in school scale (EN); BRS, brief resilience scale (FR); BSMAS, Bergen social media addiction scale (FR); BVQr, Olweus revised bully and victim questionnaire (FR); FTP, future time perspective scale (FR); GEA, global evaluation acne (FR); HEAS, Hogg eco-anxiety scale (EN); IDPSQ-14, Quebec health department's psychological distress index (FR); IGCS, Internet Gaming Cognitions Scale (FR); IPAQ, international physical activity questionnaire (FR); ISI-3, Insomnia severity index – brief version (FR); MSPSS, multidimensional scale of perceived social support (FR); PANALE, positive and negative adolescent life experiences scale (EN); PedsQL, pediatric quality of life inventory (FR); PHQ-9, patient health questionnaire for depression severity (FR); RSES, rosenberg self-esteem scale (EN); SHS, subjective happiness scale (FR); SBI, School burnout inventory (FR); WHO-5, World Health Organization well-being index. When appropriate, bilingual experts in epidemiology performed English (EN) to French (FR) translations using a forward-backward approach.

### Qualitative study population, recruitment and procedures

The quantitative findings of the cohort study were complemented and enriched by a cross-sectional qualitative study conducted between October 2023 and March 2024, focusing specifically on socioeconomically disadvantaged populations. This component, known as the PIRA project (Pandemic and Inequalities: Analysis of the Impact on Adolescents), was carried out by an external team of experts commissioned for this purpose, under a mandate assigned to the Observatory of Families, affiliated with the Institute of Sociological Research in Geneva.

Eligibility criteria included being 10 to 14 years old during the acute phase of the pandemic, and either reporting negative impacts from the pandemic or living in a low-income family, a household experiencing family tensions or conflicts, or having a family member facing health issues or disabilities.^53^ Recruitment took place in facilities offering recreational activities and individual and group support for young people, in various neighborhoods and municipalities of the canton of Geneva, as well as in municipal social services and consultation centers frequented by the parents of the targeted adolescents. Contact was established through a playful activity that helped initiate discussion and assess the presence of at least one vulnerability criterion, which led to proposing participation in the study. Semi-structured qualitative interviews were conducted to gather first-hand accounts and gain a deeper understanding of how young people experienced the period from the initial lockdown to the lifting of all restrictions, as well as any challenges they faced up to the time of the interview.

An additional component of this qualitative study involved focus groups with frontline professionals who have been working with adolescents aged 10 to 18 since 2020, including teachers, specialized educators, psychologists, and social workers. These professionals were invited to share their perspectives on the impact of the pandemic on socially vulnerable adolescents. All interviews and focus groups were recorded, then fully transcribed and anonymized before being thematically analyzed.

The PIRA study received approval from the Geneva Cantonal Commission for Research Ethics following an amendment submitted as part of the SEROCoV-KIDS study (N° 2021-01973). All participants provided written informed consent.

### Patient and public involvement

While patients and the public were not directly involved in the design, conduct, reporting, or dissemination plans of the SEROCoV-KIDS study, we included open-ended questions at the end of each survey, asking participants to provide comments or suggest topics for future questionnaires. An interactive webinar was organized in February 2022, offering participants and the wider public the opportunity to ask questions to experts. The webinar is available for replay (https://www.youtube.com/watch?v=kP6PelXh1LM). Our group sends newsletters three to four times a year and maintains a website to keep participants and the general public informed about our studies. Our website features a news feed where study findings are presented in plain language (https://www.specchio-hub.ch/actualites). We also offer phone and email support for technical or study-related questions to encourage ongoing engagement.

## Findings to date

### Cohort characteristics

A total of 2199 children and adolescents from 1340 households were enrolled in the SEROCoV-KIDS cohort, 2048 in the population-based sample and 151 in the clinical sample (Figure 1). The participation rate differed based on prior participation in seroprevalence studies (42.1% among previous participants and 9.1% among newly selected ones). Among new participants, older children and adolescents had higher participation rates (10.5% for ages 6–13 and 9.5% for ages 14–17) compared to children aged 1–5 years (7.1%). At enrollment, participants from the population-based sample were aged between 6 months and 17 years (mean: 9.8 years, standard deviation [SD]: 4.3), with a balanced sex distribution (male: 49.8%; female: 50.1%; other: 0.1%; Table 3). Most children were born in Switzerland (88.2%), had at least one parent with a tertiary education (83.8%) and lived in a household with a good or very good financial situation (81.5%). In the clinical sample, the mean age was 8.7 years (SD 4.8), and 46.4% were females. Children were mostly Swiss born (81.0%) and lived in households with highly educated parents (73.7%) and a good or very good financial situation (67.2%).

**Table 3.**
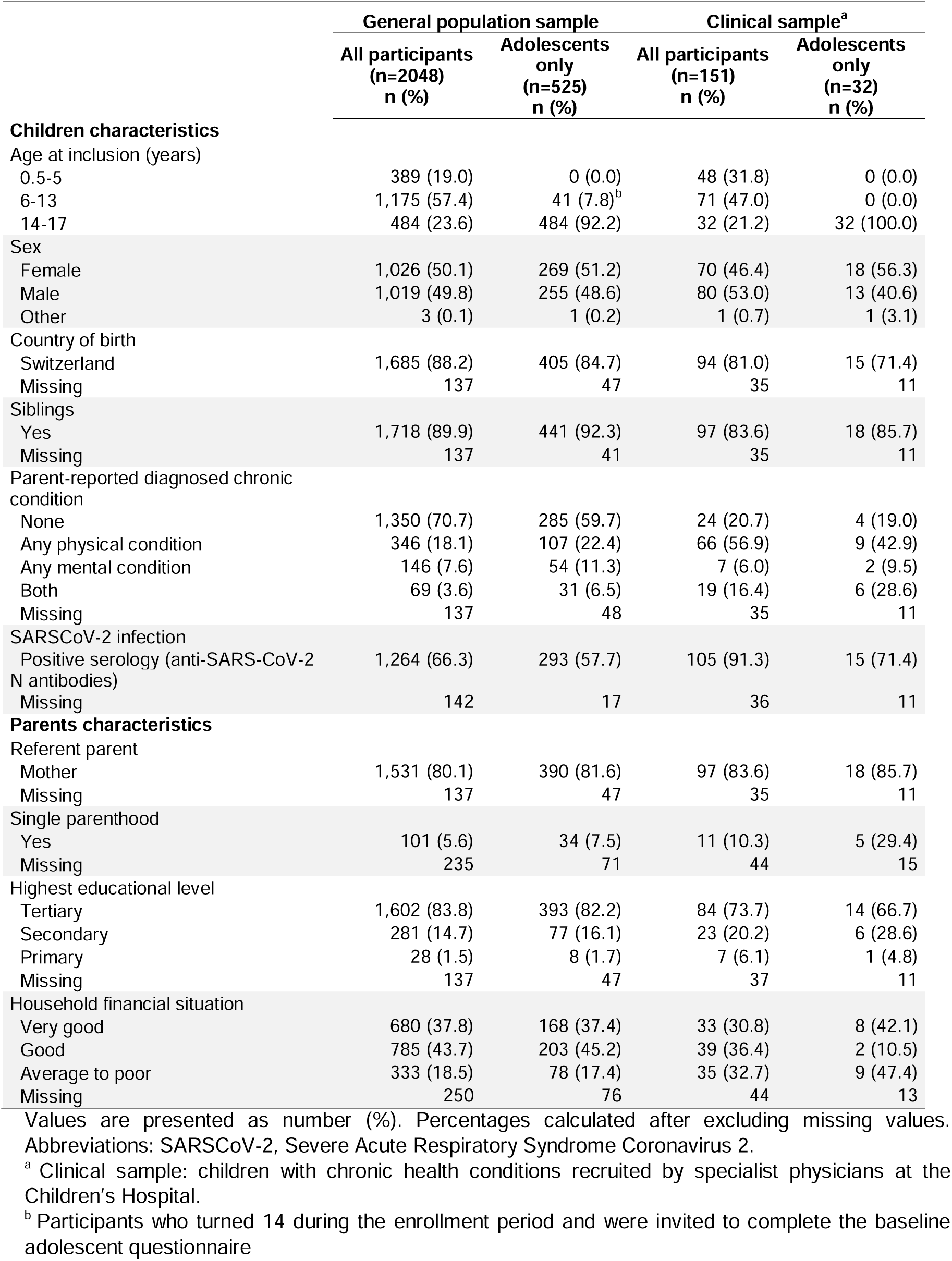
Baseline characteristics of the 2199 children included in the SEROCoV-KIDS cohort (Dec, 2021 - Dec, 2022)

### Attrition

Overall, questionnaire completion rates up to 2025 were 92.2% at baseline (2027/2199), followed by 76.9% (1692/2199), 66.8% (1464/2193), 60.8% (1302/2140), 55.6% (1190/2140), and 51.0% (1089/2134) at subsequent assessments, after excluding study withdrawals (Table 4). For adolescent questionnaires, completion rates were 80.4% at baseline (448/557), and 61.6% (343/557), 63.7% (354/556), 52.4% (311/593), 49.9% (296/593), and 46.8% (276/590) at the following assessments (variations in the denominators are due to study withdrawals and the inclusion of adolescents who turned 14). Non-response was more frequent for older, non-Swiss children, and those from single-parent families or households with average-to-poor financial situations (Tables 4, 5, 6). As of fall 2025, a total of 70 (3.2%) parents and 31 adolescents (5.6%) had withdrawn from the study.

**Table 4.**
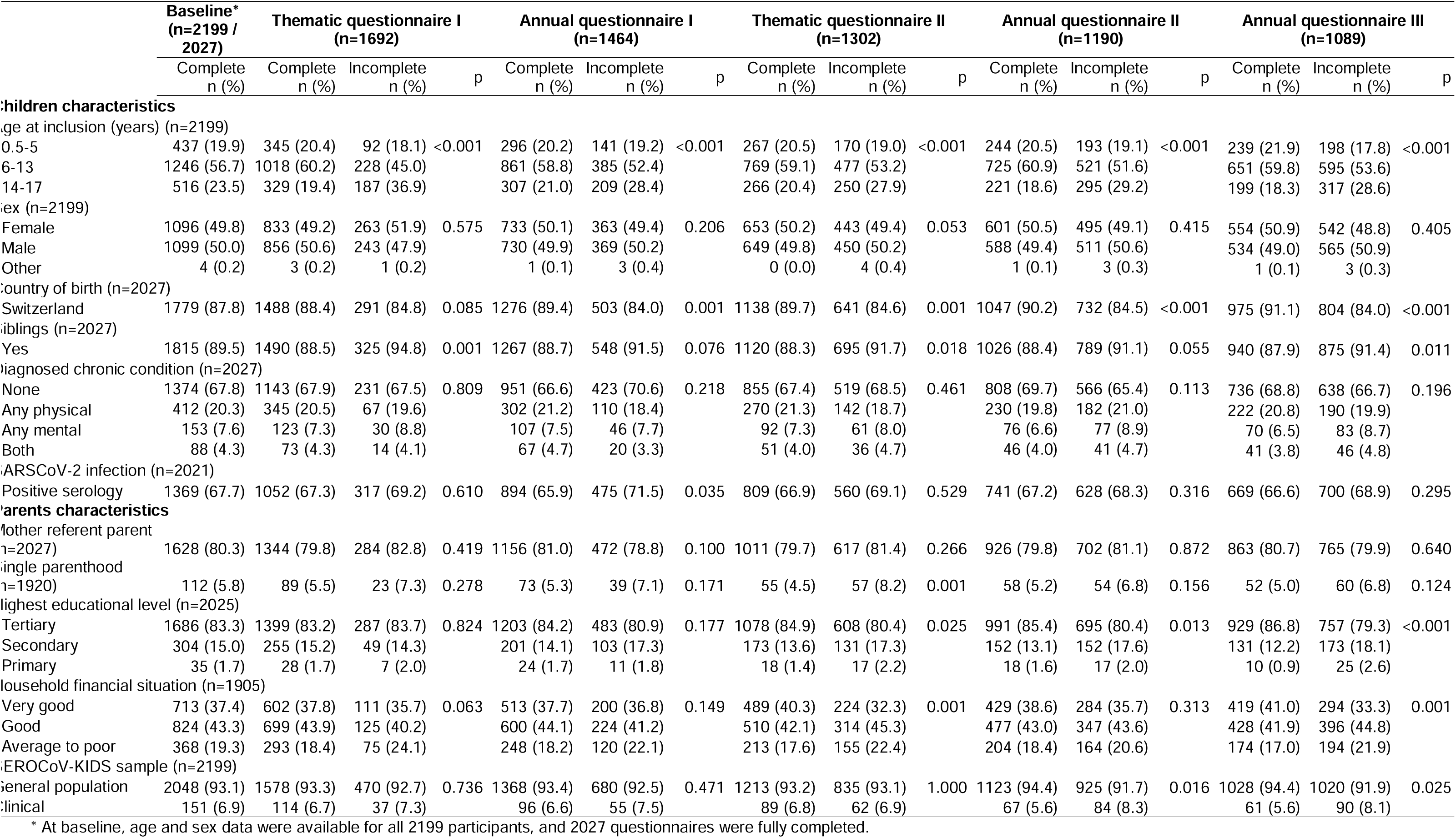
Characteristics of overall cohort participants by response to follow-up questionnaires.

**Table 5.**
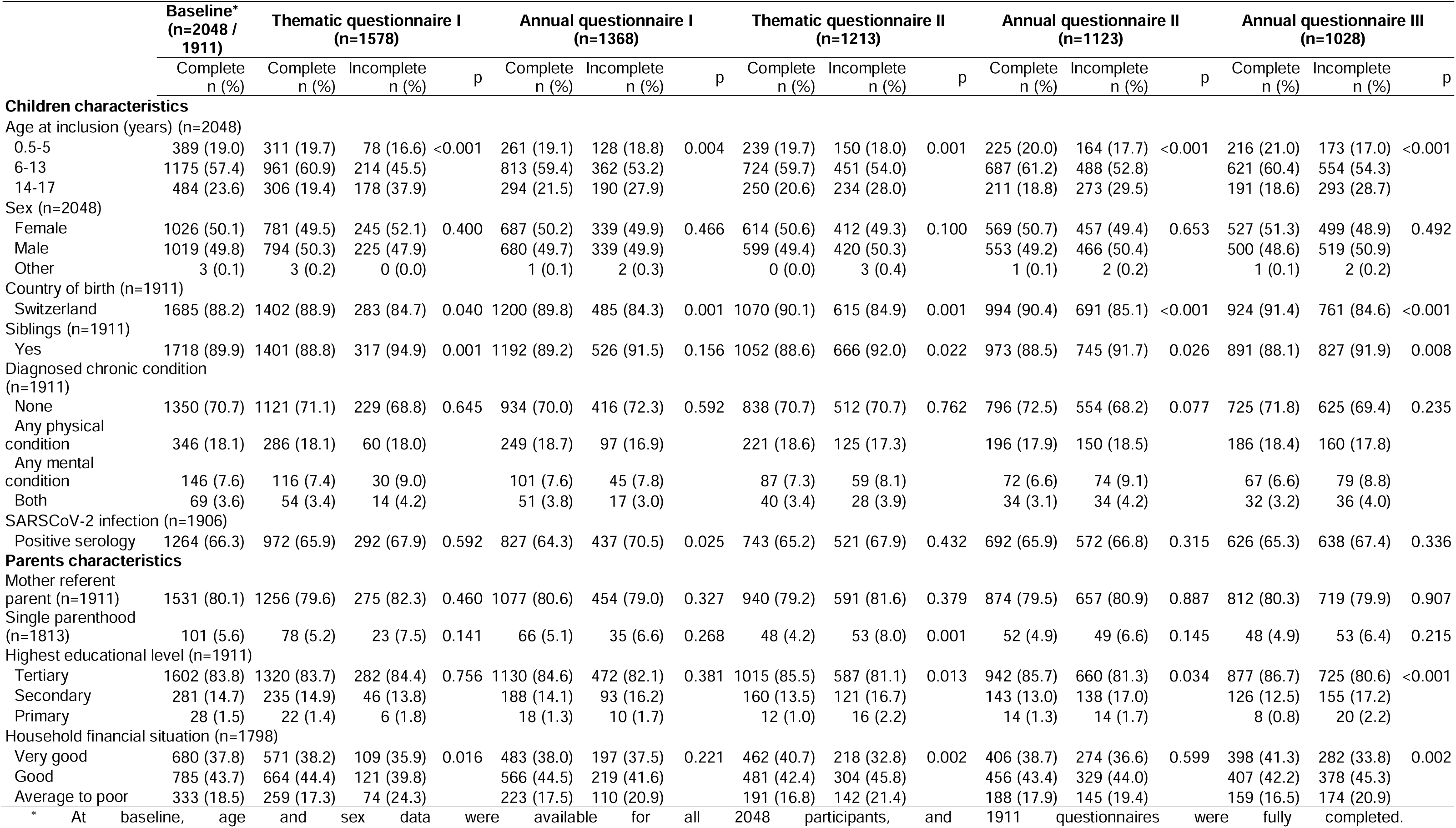
Characteristics of participants from the general population sample by response to follow-up questionnaires.

**Table 6.**
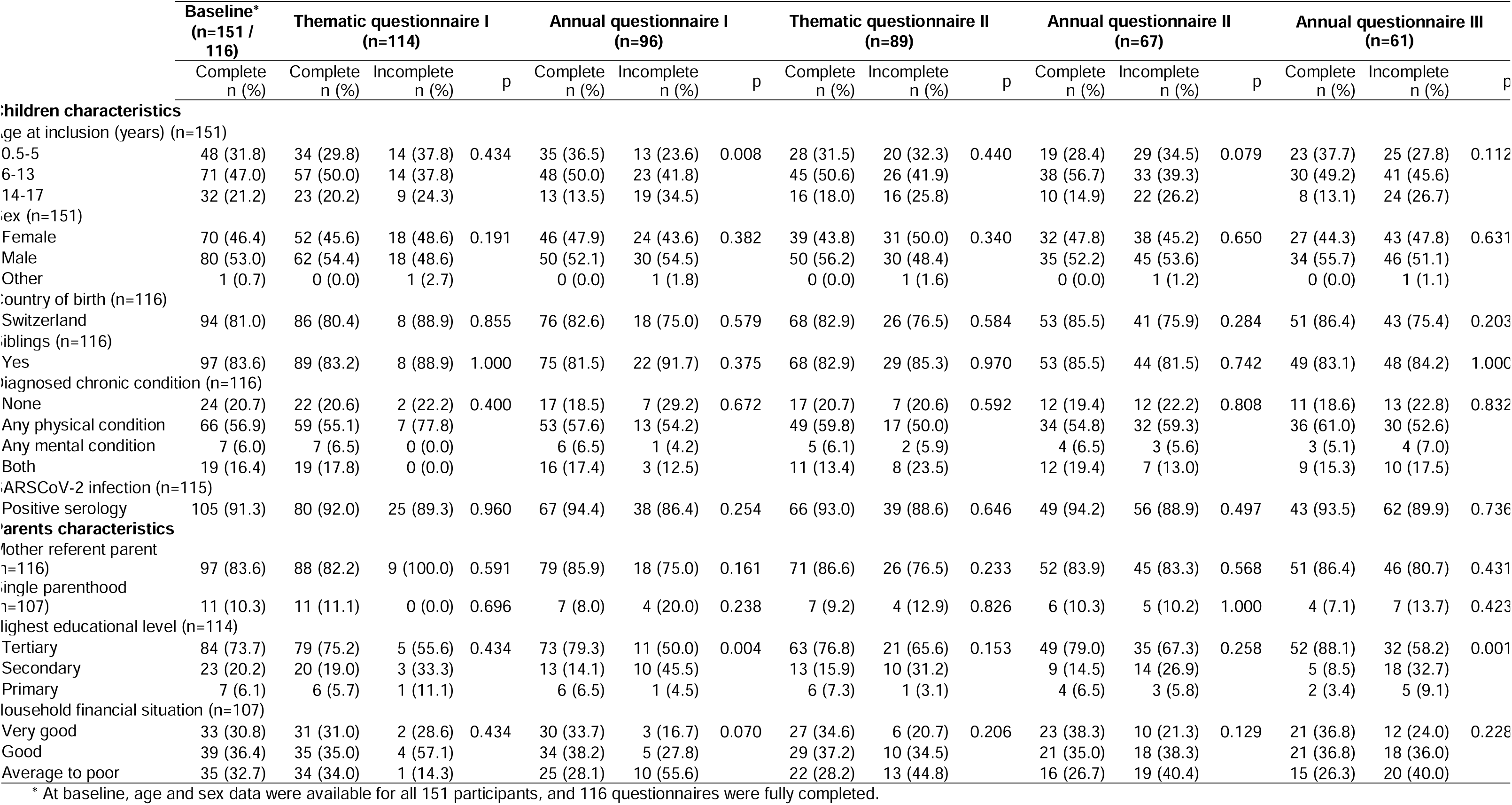
Characteristics of participants from the clinical sample (children with chronic health conditions) by response to follow-up questionnaires.

### Prevalence of anti-SARS-CoV-2 antibodies and the health of children and adolescents with prior SARS-CoV-2 infection

The baseline visit coincided with the widespread circulation of the Omicron variant, which led to a high infection rate among the pediatric population. As a result, the majority of children and adolescents tested seropositive for anti-SARS-CoV-2 N antibodies, reaching 66.3% in the population-based sample recruited between December 2021 and June 2022 and 91.3% in the clinical sample recruited between June and December 2022 (Tables 5, 6). Among them, 31.1% and 58.6% reported having had a positive PCR or rapid antigenic test. Two years into the pandemic, a substantial number of children and adolescents experienced symptoms consistent with post-COVID condition as defined by the World Health Organization in March 2022, i.e., persistent symptoms lasting more than 12 weeks after a confirmed SARS-CoV-2 infection and without an alternative diagnosis.^54^ In the population-based sample, the overall prevalence of pediatric post-COVID condition was estimated to be 4.1% (95% confidence interval [CI] 1.1–7.3), with adolescents showing a significantly higher prevalence (8.3%, 95% CI 3.5–13.5).^55^ The main risk factors identified for post-COVID condition included older age, disadvantaged socioeconomic conditions, and the presence of a chronic health condition, particularly asthma. We further explored a potential pathophysiological pathway linking acute SARS-CoV-2 infection to persistent symptoms. Our findings suggest that SARS-CoV-2 infection induces the production of autoantibodies against apolipoprotein A-1 (AAA1), which are associated with short-term symptom persistence in children, corroborating previous observations in adults.^56^

### Health and psychosocial consequences of the pandemic in the general pediatric population

In 2021-2022, at baseline, four out of five children had not been impacted by the pandemic, or only minimally, and displayed low levels of mental health difficulties throughout the 2022-2025 period.^57,58^ This overall reassuring finding suggests that Switzerland’s relatively light restrictions and broad social benefits helped mitigate the pandemic’s health impact on children. Approximately 8% of participants reported a positive pandemic impact. Their baseline mental health and evolution over time were comparable to those of participants who experienced no impact. Conversely, depending on the measurement tool, 8 to 13% of children and their households experienced a pronounced negative impact, mainly due to disruptions in daily routines and reduced access to social and family support.^57,58^ They exhibited poorer health-related quality of life (HRQoL) and mental health in 2022.^57,58^ Although mental health improved in this group between 2022 and 2025, it remained poorer than among children who were not impacted.^58^ While these findings are reassuring overall, they underscore that the pandemic’s effects on youth well-being persisted for years, even after most restrictions had been lifted.

Since many families reported challenges in balancing work and childcare early in the pandemic, we investigated whether parents’ work-to-family conflict was associated with trajectories of mental health for both parents and children between 2022 and 2024.^59^ Higher levels of conflict were consistently associated with worse anxiety and depressive symptoms in parents, as well as with internalizing and externalizing difficulties in children.

We further explored specific aspects of the mental health of adolescents. This developmental stage is characterized by profound physical, psychological, and social changes that significantly influence the transition into adulthood.^60^ Each generation of adolescents faces unique contextual challenges, such as the pandemic and related restrictions, adding complexity to their developmental trajectory. In 2022, we observed that 14% of adolescents in the population-based sample reported suicidal ideation in the previous year, a rate similar to global pre-pandemic levels.^61^ Key risk factors included psychological distress, low self-esteem, identification with a sexual minority, bullying, excessive screen time, and a severe pandemic impact. We also examined the psychological burden of acne, finding that adolescents with lower acne-specific quality of life experienced greater distress, lower self-esteem, and reduced resilience.^62^ These effects were mitigated by physical activity and social support, but worsened by screen time and social media addiction. Lastly, in another study we reported that one in six adolescents felt limited in their future perspectives, and this was particularly true for those with mental health difficulties, limited social support, academic struggles, sexual minority identity and excessive screen time. Major concerns were centered on failure, education, and climate change.^63^ In adolescents, many risk factors identified are interrelated and contribute to multiple mental health challenges, underscoring the need for integrated, multifaceted approaches to prevention and intervention. Conversely, strong parent-child relationships, social support, physical activity, and high self-esteem emerged as key protective factors across various domains of mental health.^61–63^

Health behaviors are essential to the physical and mental health of children and adolescents.^64^ Prolonged pandemic control measures, such as remote schooling, limitation of in-person interactions and restricted access to recreational facilities, disrupted the health behaviors of young people. In Switzerland, however, these restrictions were comparatively milder than in many neighboring countries.^65^ Our findings show that, two years into the pandemic, 20% to 30% of children failed to meet health behavior recommendations for physical activity, screen time and sleep duration.^66^ For most children, physical activity (72.9%), time spent in green spaces (72.2%), and sleep quality (93.1%) remained unchanged throughout the pandemic, while over half (51.9%) reported increased screen time. This increase in screen time is concerning given the observed association between screen time and lower quality of life,^62,67^ psychological distress^61^ or future perspectives^63^ among preadolescents and adolescents.

### Health and psychosocial consequences of the pandemic in children with chronic health conditions

Children living with chronic health conditions or having special health care needs face greater challenges to mental health and quality of life compared to the general pediatric population.^68–71^ Our findings showed that, although these children were not at higher risk of SARS-CoV-2 infection compared to their healthy peers, they were more likely to suffer from persistent symptoms^55^ and to report a severe pandemic impact.^57,72^ After pandemic-related restrictions were lifted, children with special healthcare needs had poorer well-being than healthy children, with a gradient according to the complexity of their condition.^72^ Children with chronic conditions also experienced greater difficulties in physical and social functioning and externalizing behaviors, but reported similar levels of prosocial behavior, social support, and extracurricular participation as their healthy peers. Those with complex chronic conditions were at particularly high risk for poor well-being across physical, psychological, and social domains. Even after restrictions ended, only a few well-being aspects improved, while many others worsened.^72^ These results highlight the need for long-term follow-up of post-pandemic well-being and targeted interventions to support these children.

### Health and psychosocial consequences of the pandemic in socioeconomically disadvantaged populations

Several of our studies have highlighted how the pandemic exacerbated existing social inequalities in health. Socioeconomic and family disadvantage was consistently associated with a more severe pandemic impact^57,73^ and greater psychosocial difficulties (having few close friends, school difficulties, no engagement in extracurricular activities).^74,75^ Disadvantaged children were also more likely to engage in unhealthy behaviors (not meeting sleep duration guidelines, limited physical activity, smoking, soda consumption, low adherence to COVID-19 vaccination guidelines),^66,74,76^ and were more frequently affected by a range of unfavorable health outcomes, including persistent symptoms compatible with post-COVID condition,^55^ poor health-related quality of life,^74,77^ internalizing and externalizing difficulties,^77^ poorer well-being and higher anxiety at school.^75^ Our results also suggest that certain psychosocial factors, such as a good parent-child relationship or social support, can act as protective factors for poor outcomes.^61,62,74^

Disadvantaged children showed a milder mental health response to the negative impact of the pandemic than advantaged children, and mental health difficulties decreased over time in both groups, regardless of the financial situation of their households.^73^ This illustrates the complexity of the pandemic’s social impact and suggests that, while disadvantaged children were more vulnerable to pandemic-related burdens, many demonstrated coping resources and notable resilience.

The qualitative PIRA study further illuminated this complexity by capturing the experiences of 17 adolescents and 20 professionals working with adolescents.^53^ It highlighted the differentiated effects of the health crisis on young people, especially the conflicting expectations they faced across their school, family, and social roles, which they had to adopt, abandon, and relearn. The shift to remote learning exacerbated educational inequalities, making reintegration more challenging for vulnerable youth, sometimes leading to disengagement. Family life had contrasting effects: while some households experienced strengthening bonds, others, particularly those in precarious conditions, faced increased tensions. Social isolation, especially among certain groups such as girls, was partly offset by intensified online interactions, with mixed consequences. The return to normalcy revealed and exacerbated social and psychological stress. This study component showed that the most vulnerable youth accumulated difficulties during both the acute phase of the pandemic and the return to normalcy, widening social disparities with potentially lasting effects on their trajectories. It emphasized the need to analyze these experiences within relational dynamics between peers and adults, reflecting the quantitative findings of the cohort.

## Strengths and limitations

The SEROCoV-KIDS study is one of the few pediatric cohorts established during the pandemic. Its main strengths include its population-based design with random sampling, its large sample size, the broad age range covered, its longitudinal follow-up, and the use of a combination of quantitative and qualitative approaches, all of which enhance the external validity and relevance of the findings. However, several limitations should be considered when interpreting our findings. First, the lack of individual-level pre- and early-pandemic data limits direct comparisons of changes in children’s health, well-being and behaviors over time. Instead, the study relies on parent-reported assessments of the pandemic’s impact which may be affected by recall bias. Consequently, robust conclusions regarding temporal trends can only be drawn from repeated measures collected between 2021 and 2025. Nevertheless, these findings still provide valuable insights given the lack of a national cohort of children in Switzerland. Second, despite random sampling, participation bias led to the overrepresentation of children aged 8 to 14, because parents were often reluctant to have younger children undergo a blood draw solely for research purposes and because adolescents had to complete their own questionnaires, which might have reduced their willingness to participate. As commonly observed in epidemiological studies, highly educated families were also overrepresented in the cohort, likely reflecting greater social engagement and trust in science among more privileged groups.^78^ This may limit the generalizability of our findings and could lead to an overestimation of population-level well-being and health behaviors. Additionally, even though one-fifth of the cohort reported financial difficulties, the most disadvantaged children were likely underrepresented, potentially leading to an underestimation of adverse outcomes in populations facing the greatest pandemic-related challenges. Finally, despite various incentives to maintain participation, the study experienced “COVID fatigue”, resulting in substantial attrition over time.

## Collaboration

We welcome collaborative projects, and researchers may apply for access to the SEROCoV-KIDS biobank and cohort data, subject to authorization from our scientific board and ethics approval for their study. Requests for study questionnaires and data access can be submitted to.

## Conclusions and future plans

The SEROCoV-KIDS cohort demonstrates that sustained, population-based pediatric research is essential for monitoring and understanding how public policies and major societal events shape child and adolescent health, and for informing evidence-based decision-making. As a uniquely large pediatric cohort study in Switzerland, SEROCoV-KIDS strengthens the evidence base on how the pandemic and its aftermath have affected children and adolescents, revealing both resilience and persistent inequalities. Its findings underscore the need to invest in long-term, population-based cohort infrastructures capable of generating timely, multidimensional data that integrate biological, psychological, and social dimensions. Such sustained monitoring systems are essential for detecting early warning signals, evaluating interventions, and designing preventive strategies that promote health equity and well-being. More than a scientific imperative, this investment is a public responsibility. SEROCoV-KIDS provides a model for anticipatory, evidence-informed governance that shifts societies from crisis response toward building environments that protect, empower, and support future generations.

## Acknowledgements

We are grateful to the staff of the Unit of Population Epidemiology, as well as to all participants whose contributions were invaluable to the study. We also thank the members of the sérothèque from the Division of Laboratory Medicine at HUG for sample preparation and serological testing.

## Collaborators

The SEROCoV-KIDS Study Group include the following collaborators (including the members listed as authors): Deborah Amrein, Andrew S. Azman, Antoine Bal, Rémy P. Barbe, Hélène Baysson, Julie Berthelot, Patrick Bleich, Livia Boehm, Aminata R. Bouhet, Nicolas Bovio, Gaëlle Bryand-Rumley, Viola Bucolli, Prune Collombet, Vladimir Davidovic, Paola D’Ippolito, Richard Dubos, Roxane Dumont, Nacira El Merjani, Natalia Fernandez Clares, Natalie Francioli, Clément Graindorge, Idris Guessous, Munire Hagose, Séverine Harnal, Julien Lamour, Pierre Lescuyer, Arnaud G. L’Huillier, Andrea Loizeau, Elsa Lorthe, Claire Mariano, Chantal Martinez, Shannon Mechoullam, Mayssam Nehme, Natacha Noël, Francesco Pennacchio, Javier Perez-Saez, Klara M. Posfay-Barbe, Géraldine Poulain, Caroline Pugin, Nick Pullen, Viviane Richard, Deborah Rochat, Cyril Sahyoun, Irine Sakvarelidze, Khadija Samir, Manuel Schibler, Stephanie Schrempft, Jessica Rizzo, Silvia Stringhini, Stéphanie Testini, Deborah Urrutia Rivas, Charlotte Verolet, Jennifer Villers, Guillemette Violot, Nicolas Vuillemier, María-Eugenia Zaballa.

## CRediT authorship contribution statement

Elsa Lorthe: Conceptualization, Funding acquisition, Investigation, Validation, Writing – original draft. Andrea Loizeau: Funding acquisition, Investigation, Project administration, Writing – original draft. Viviane Richard: Investigation, Formal analysis, Data curation, Validation, Writing – review & editing. Roxane Dumont: Investigation, Data curation, Validation, Writing – review & editing. María-Eugenia Zaballa: Investigation, Writing – review & editing. Francesco Pennacchio: Software, Data curation, Validation, Writing – review & editing. Julien Lamour: Software, Formal analysis, Data curation, Validation, Writing – review & editing. Arnaud G L’Huillier: Supervision, Writing – review & editing. Hélène Baysson: Conceptualization, Funding acquisition, Investigation, Writing – review & editing. Natalia Fernandez Clares: Investigation, Project administration, Writing – review & editing. Nicolas Bovio: Investigation, Validation, Writing – review & editing. Mayssam Nehme: Supervision, Writing – review & editing. Pierre Lescuyer: Supervision, Writing – review & editing. Nicolas Vuilleumier: Supervision, Writing – review & editing. Klara M. Posfay-Barbe: Conceptualization, Funding acquisition, Supervision, Writing – review & editing. Rémy P. Barbe: Conceptualization, Funding acquisition, Supervision, Writing – review & editing. Idris Guessous: Conceptualization, Funding acquisition, Investigation, Supervision, Writing – review & editing. Silvia Stringhini: Conceptualization, Funding acquisition, Project administration, Investigation, Supervision, Writing – review & editing. All authors approved the final version to be published.

## Participants consent

Adolescents from 14 years of age and parents of all participants gave informed consent to participate. Children gave oral assent to participate.

## Ethics approval

This study involves human participants and its conduct was approved by the Cantonal Research Ethics Commission of Geneva (CCER, N°2021-01973).

## Provenance and peer review

Not commissioned; externally peer reviewed.

## Data sharing statement

Researchers may apply for access to the SEROCoV-KIDS biobank and cohort data, subject to ethics approval for their study and authorization from our scientific board. Requests for study questionnaires and data access can be submitted to.

## References

1. Viner RM, Mytton OT, Bonell C, et al. Susceptibility to SARS-CoV-2 Infection Among Children and Adolescents Compared With Adults: A Systematic Review and Meta-analysis. JAMA Pediatr. 2021;175(2):143–156. doi:10.1001/jamapediatrics.2020.4573

2. CDC COVID-19 Response Team. Coronavirus Disease 2019 in Children - United States, February 12-April 2, 2020. MMWR Morb Mortal Wkly Rep. 2020;69(14):422–426. doi:10.15585/mmwr.mm6914e4

3. Wiersinga WJ, Rhodes A, Cheng AC, Peacock SJ, Prescott HC. Pathophysiology, Transmission, Diagnosis, and Treatment of Coronavirus Disease 2019 (COVID-19): A Review. JAMA. 2020;324(8):782–793. doi:10.1001/jama.2020.12839

4. Docherty AB, Harrison EM, Green CA, et al. Features of 20 133 UK patients in hospital with covid-19 using the ISARIC WHO Clinical Characterisation Protocol: prospective observational cohort study. BMJ. 2020;369:m1985. doi:10.1136/bmj.m1985

5. Riccardo F, Ajelli M, Andrianou XD, et al. Epidemiological characteristics of COVID-19 cases and estimates of the reproductive numbers 1 month into the epidemic, Italy, 28 January to 31 March 2020. Eurosurveillance. 2020;25(49):2000790. doi:10.2807/1560-7917.ES.2020.25.49.2000790

6. Perez-Saez J, Bellon M, Lessler J, et al. Evolving infectious disease dynamics shape school-based intervention effectiveness. Nat Commun. 2025;16(1):6597. doi:10.1038/s41467-025-61925-5

7. Lorthe E, Bellon M, Michielin G, et al. Epidemiological, Virological and Serological Investigation into a SARS-CoV-2 Outbreak (Alpha Variant) in a Primary School: A Prospective Longitudinal Study.; 2021:2021.10.26.21265509. doi:10.1101/2021.10.26.21265509

8. Lorthe E, Bellon M, Berthelot J, et al. A SARS-CoV-2 omicron (B.1.1.529) variant outbreak in a primary school in Geneva, Switzerland. The Lancet Infectious Diseases. 2022;0(0). doi:10.1016/S1473-3099(22)00267-5

9. Markelz A, Zirnhelt Z, Morris K, et al. Association between age of paediatric index cases and household SARS-CoV-2 transmission. Epidemiology & Infection. 2024;152:e145. doi:10.1017/S0950268824000918

10. Cordery R, Reeves L, Zhou J, et al. Transmission of SARS-CoV-2 by children to contacts in schools and households: a prospective cohort and environmental sampling study in London. The Lancet Microbe. 2022;3(11):e814–e823. doi:10.1016/S2666-5247(22)00124-0

11. Uthman OA, Lyngse FP, Anjorin S, et al. Susceptibility and infectiousness of SARS-CoV-2 in children versus adults, by variant (wild-type, alpha, delta): A systematic review and meta-analysis of household contact studies. PLOS ONE. 2024;19(9):e0306740. doi:10.1371/journal.pone.0306740

12. Galmiche S, Charmet T, Rakover A, et al. Risk of SARS-CoV-2 Infection Among Households With Children in France, 2020-2022. JAMA Netw Open. 2023;6(9):e2334084. doi:10.1001/jamanetworkopen.2023.34084

13. Mensah AA, Campbell H, Stowe J, et al. Risk of SARS-CoV-2 reinfections in children: a prospective national surveillance study between January, 2020, and July, 2021, in England. The Lancet Child & Adolescent Health. 2022;6(6):384–392. doi:10.1016/S2352-4642(22)00059-1

14. Kompaniyets L, Agathis NT, Nelson JM, et al. Underlying Medical Conditions Associated With Severe COVID-19 Illness Among Children. JAMA Netw Open. 2021;4(6):e2111182. doi:10.1001/jamanetworkopen.2021.11182

15. Ward JL, Harwood R, Kenny S, et al. Pediatric Hospitalizations and ICU Admissions Due to COVID-19 and Pediatric Inflammatory Multisystem Syndrome Temporally Associated With SARS-CoV-2 in England. JAMA Pediatrics. 2023;177(9):947–955. doi:10.1001/jamapediatrics.2023.2357

16. Harwood R, Yan H, Talawila Da Camara N, et al. Which children and young people are at higher risk of severe disease and death after hospitalisation with SARS-CoV-2 infection in children and young people: A systematic review and individual patient meta-analysis. EClinicalMedicine. 2022;44:101287. doi:10.1016/j.eclinm.2022.101287

17. Ravens-Sieberer U, Kaman A, Erhart M, Devine J, Schlack R, Otto C. Impact of the COVID-19 pandemic on quality of life and mental health in children and adolescents in Germany. Eur Child Adolesc Psychiatry. 2022;31(6):879–889. doi:10.1007/s00787-021-01726-5

18. Madigan S, Eirich R, Pador P, McArthur BA, Neville RD. Assessment of Changes in Child and Adolescent Screen Time During the COVID-19 Pandemic: A Systematic Review and Meta-analysis. JAMA Pediatr. 2022;176(12):1188–1198. doi:10.1001/jamapediatrics.2022.4116

19. Ludwig-Walz H, Siemens W, Heinisch S, Dannheim I, Loss J, Bujard M. How the COVID-19 pandemic and related school closures reduce physical activity among children and adolescents in the WHO European Region: a systematic review and meta-analysis. Int J Behav Nutr Phys Act. 2023;20(1):149. doi:10.1186/s12966-023-01542-x

20. Prime H, Wade M, Browne DT. Risk and resilience in family well-being during the COVID-19 pandemic. Am Psychol. 2020;75(5):631–643. doi:10.1037/amp0000660

21. Garagiola ER, Lam Q, Wachsmuth LS, et al. Adolescent Resilience during the COVID-19 Pandemic: A Review of the Impact of the Pandemic on Developmental Milestones. Behavioral Sciences. 2022;12(7):220. doi:10.3390/bs12070220

22. Foster S, Estévez-Lamorte N, Walitza S, Dzemaili S, Mohler-Kuo M. Perceived stress, coping strategies, and mental health status among adolescents during the COVID-19 pandemic in Switzerland: a longitudinal study. Eur Child Adolesc Psychiatry. 2023;32(6):937–949. doi:10.1007/s00787-022-02119-y

23. Coker TR, Cheng TL, Ybarra M. Addressing the Long-term Effects of the COVID-19 Pandemic on Children and Families: A Report From the National Academies of Sciences, Engineering, and Medicine. JAMA. 2023;329(13):1055–1056. doi:10.1001/jama.2023.4371

24. Rider EA, Ansari E, Varrin PH, Sparrow J. Mental health and wellbeing of children and adolescents during the covid-19 pandemic. BMJ. 2021;374:n1730. doi:10.1136/bmj.n1730

25. Robinson E, Sutin AR, Daly M, Jones A. A systematic review and meta-analysis of longitudinal cohort studies comparing mental health before versus during the COVID-19 pandemic in 2020. Journal of Affective Disorders. 2022;296:567–576. doi:10.1016/j.jad.2021.09.098

26. Sun Y, Wu Y, Fan S, et al. Comparison of mental health symptoms before and during the covid-19 pandemic: evidence from a systematic review and meta-analysis of 134 cohorts. BMJ. 2023;380:e074224. doi:10.1136/bmj-2022-074224

27. Essau CA, de la Torre-Luque A. Adolescent psychopathological profiles and the outcome of the COVID-19 pandemic: Longitudinal findings from the UK Millennium Cohort Study. Progress in Neuro-Psychopharmacology and Biological Psychiatry. 2021;110:110330. doi:10.1016/j.pnpbp.2021.110330

28. Ludwig-Walz H, Dannheim I, Pfadenhauer LM, Fegert JM, Bujard M. Anxiety increased among children and adolescents during pandemic-related school closures in Europe: a systematic review and meta-analysis. Child Adolesc Psychiatry Ment Health. 2023;17(1):74. doi:10.1186/s13034-023-00612-z

29. Ludwig-Walz H, Dannheim I, Pfadenhauer LM, Fegert JM, Bujard M. Increase of depression among children and adolescents after the onset of the COVID-19 pandemic in Europe: a systematic review and meta-analysis. Child Adolesc Psychiatry Ment Health. 2022;16(1):109. doi:10.1186/s13034-022-00546-y

30. Ma L, Mazidi M, Li K, et al. Prevalence of mental health problems among children and adolescents during the COVID-19 pandemic: A systematic review and meta-analysis. J Affect Disord. 2021;293:78–89. doi:10.1016/j.jad.2021.06.021

31. Racine N, McArthur BA, Cooke JE, Eirich R, Zhu J, Madigan S. Global Prevalence of Depressive and Anxiety Symptoms in Children and Adolescents During COVID-19: A Meta-analysis. JAMA Pediatr. 2021;175(11):1142–1150. doi:10.1001/jamapediatrics.2021.2482

32. Madigan S, Racine N, Vaillancourt T, et al. Changes in Depression and Anxiety Among Children and Adolescents From Before to During the COVID-19 Pandemic: A Systematic Review and Meta-analysis. JAMA Pediatr. 2023;177(6):567–581. doi:10.1001/jamapediatrics.2023.0846

33. O’Connor M, Olsson CA, Lange K, et al. Progressing “Positive Epidemiology”: A Cross-national Analysis of Adolescents’ Positive Mental Health and Outcomes During the COVID-19 Pandemic. Epidemiology. 2025;36(1):28–39. doi:10.1097/EDE.0000000000001798

34. Campione-Barr N, Skinner A, Moeller K, Cui L, Kealy C, Cookston J. The role of family relationships on adolescents’ development and adjustment during the COVID-19 pandemic: A systematic review. Journal of Research on Adolescence. 2025;35(1):e12969. doi:10.1111/jora.12969

35. Seligman B, Ferranna M, Bloom DE. Social determinants of mortality from COVID-19: A simulation study using NHANES. PLOS Medicine. 2021;18(1):e1003490. doi:10.1371/journal.pmed.1003490

36. Dragano N, Dortmann O, Timm J, et al. Association of Household Deprivation, Comorbidities, and COVID-19 Hospitalization in Children in Germany, January 2020 to July 2021. JAMA Network Open. 2022;5(10):e2234319. doi:10.1001/jamanetworkopen.2022.34319

37. Mannheim J, Konda S, Logan LK. Racial, ethnic and socioeconomic disparities in SARS-CoV-2 infection amongst children. Paediatr Perinat Epidemiol. 2022;36(3):337–346. doi:10.1111/ppe.12865

38. Holmberg V, Salmi H, Kattainen S, et al. Association between first language and SARS-CoV-2 infection rates, hospitalization, intensive care admissions and death in Finland: a population-based observational cohort study. Clin Microbiol Infect. 2022;28(1):107–113. doi:10.1016/j.cmi.2021.08.022

39. Reiss F. Socioeconomic inequalities and mental health problems in children and adolescents: a systematic review. Soc Sci Med. 2013;90:24–31. doi:10.1016/j.socscimed.2013.04.026

40. Xiao Y, Yip PSF, Pathak J, Mann JJ. Association of Social Determinants of Health and Vaccinations With Child Mental Health During the COVID-19 Pandemic in the US. JAMA Psychiatry. 2022;79(6):610–621. doi:10.1001/jamapsychiatry.2022.0818

41. Xiao Y, Brown TT, Snowden LR, Chow JCC, Mann JJ. COVID-19 Policies, Pandemic Disruptions, and Changes in Child Mental Health and Sleep in the United States. JAMA Netw Open. 2023;6(3):e232716. doi:10.1001/jamanetworkopen.2023.2716

42. O’Connor M, Lange K, Downes M, et al. Socio-economic disparities in the psychosocial and economic impacts of the COVID-19 pandemic on children and young people in Australia. Journal of Paediatrics and Child Health. 2025;61(2):267–276. doi:10.1111/jpc.16737

43. Gianfredi V, Scarioni S, Marchesi L, et al. The impact of the COVID-19 pandemic on academic performance among developmental age students: a systematic review with meta-analysis. Ann Ig. 2025;37(1):49–73. doi:10.7416/ai.2024.2647

44. Cortés-Albornoz MC, Ramírez-Guerrero S, García-Guáqueta DP, Vélez-Van-Meerbeke A, Talero-Gutiérrez C. Effects of remote learning during COVID-19 lockdown on children’s learning abilities and school performance: A systematic review. Int J Educ Dev. 2023;101:102835. doi:10.1016/j.ijedudev.2023.102835

45. Betthäuser BA, Bach-Mortensen AM, Engzell P. A systematic review and meta-analysis of the evidence on learning during the COVID-19 pandemic. Nat Hum Behav. 2023;7(3):375–385. doi:10.1038/s41562-022-01506-4

46. Schiera M, Faraci F, Mannino G, Vantaggiato L. The impact of the pandemic on psychophysical well-being and quality of learning in the growth of adolescents (aged 11-13): a systematic review of the literature with a PRISMA method. Front Psychol. 2024;15:1384388. doi:10.3389/fpsyg.2024.1384388

47. Stringhini S, Wisniak A, Piumatti G, et al. Seroprevalence of anti-SARS-CoV-2 IgG antibodies in Geneva, Switzerland (SEROCoV-POP): a population-based study. The Lancet. 2020;396(10247):313–319. doi:10.1016/S0140-6736(20)31304-0

48. Zaballa ME, Perez-Saez J, de Mestral C, et al. Seroprevalence of anti-SARS-CoV-2 antibodies and cross-variant neutralization capacity after the Omicron BA.2 wave in Geneva, Switzerland: a population-based study. The Lancet Regional Health - Europe. 2023;24:100547. doi:10.1016/j.lanepe.2022.100547

49. Stringhini S, Zaballa ME, Perez-Saez J, et al. Seroprevalence of anti-SARS-CoV-2 antibodies after the second pandemic peak. The Lancet Infectious Diseases. 2021;21(5):600–601. doi:10.1016/S1473-3099(21)00054-2

50. Stringhini S, Zaballa ME, Pullen N, et al. Seroprevalence of anti-SARS-CoV-2 antibodies 6 months into the vaccination campaign in Geneva, Switzerland, 1 June to 7 July 2021. Eurosurveillance. 2021;26(43):2100830. doi:10.2807/1560-7917.ES.2021.26.43.2100830

51. Stringhini S, Zaballa ME, Pullen N, et al. Large variation in anti-SARS-CoV-2 antibody prevalence among essential workers in Geneva, Switzerland. Nat Commun. 2021;12(1):3455. doi:10.1038/s41467-021-23796-4

52. Baysson H, Pennacchio F, Wisniak A, et al. Specchio-COVID19 cohort study: a longitudinal follow-up of SARS-CoV-2 serosurvey participants in the canton of Geneva, Switzerland. BMJ Open. 2022;12(1):e055515. doi:10.1136/bmjopen-2021-055515

53. Consoli L, Widmer E, Burton C. Répercussions de la pandémie de COVID-19 sur les adolescents à Genève: stress social et changements de rôles. Genève: Université de Genève Sociograph 71. Published online 2025. https://www.unige.ch/sciences-societe/socio/application/files/1017/3917/4637/Sociograph_71_COVID.pdf

54. WHO. A clinical case definition for post COVID-19 condition in children and adolescents by expert consensus, 16 February 2023. February 16, 2023. Accessed October 1, 2025. https://www.who.int/publications/i/item/WHO-2019-nCoV-Post-COVID-19-condition-CA-Clinical-case-definition-2023-1

55. Dumont R, Richard V, Lorthe E, et al. A population-based serological study of post-COVID syndrome prevalence and risk factors in children and adolescents. Nat Commun. 2022;13:7086. doi:10.1038/s41467-022-34616-8

56. Vuilleumier N, Pagano S, Lorthe E, et al. Association between SARS-CoV-2 infection and anti-apolipoprotein A-1 antibody in children. Front Immunol. 2025;16:1521299. doi:10.3389/fimmu.2025.1521299

57. Richard V, Dumont R, Lorthe E, et al. Impact of the COVID-19 pandemic on children and adolescents: determinants and association with quality of life and mental health—a cross-sectional study. Child and Adolescent Psychiatry and Mental Health. 2023;17(1):17. doi:10.1186/s13034-023-00563-5

58. Richard V, Lorthe E, Dumont R, et al. Mental health trajectories of children and adolescents up to five years after the onset of the COVID-19 pandemic: a longitudinal study. medRxiv. Preprint posted online December 2, 2025:2025.11.30.25340982. doi:10.64898/2025.11.30.25340982

59. Lorthe E, Bovio N, Uppal A, et al. Work-to-family conflict and mental health trajectories in parents and children: findings from two cohort studies. In preparation.

60. Sawyer SM, Azzopardi PS, Wickremarathne D, Patton GC. The age of adolescence. The Lancet Child & Adolescent Health. 2018;2(3):223–228. doi:10.1016/S2352-4642(18)30022-1

61. Dumont R, Lorthe E, Richard V, et al. Prevalence of and risk factors for suicidal ideation in adolescents during the COVID-19 pandemic: a cross-sectional study. Swiss Med Wkly. 2024;154:3461. doi:10.57187/s.3461

62. Dumont S, Lorthe E, Loizeau A, et al. Acne-related quality of life and mental health among adolescents: a cross-sectional analysis. Clin Exp Dermatol. 2025;50(4):795–803. doi:10.1093/ced/llae453

63. Dumont R, Lorthe E, Richard V, et al. Future time perspectives and concerns among adolescents in 2022. BMJ Paediatr Open. 2024;8(1):e002367. doi:10.1136/bmjpo-2023-002367

64. Sampasa-Kanyinga H, Colman I, Goldfield GS, et al. Combinations of physical activity, sedentary time, and sleep duration and their associations with depressive symptoms and other mental health problems in children and adolescents: a systematic review. Int J Behav Nutr Phys Act. 2020;17(1):72. doi:10.1186/s12966-020-00976-x

65. Hale T, Angrist N, Goldszmidt R, et al. A global panel database of pandemic policies (Oxford COVID-19 Government Response Tracker). Nat Hum Behav. 2021;5(4):529–538. doi:10.1038/s41562-021-01079-8

66. Richard V, Lorthe E, Dumont R, et al. Socio-demographic correlates of health behaviors of children and adolescents during the COVID-19 pandemic: a cross-sectional study. Sci Rep. 2025;15(1):30291. doi:10.1038/s41598-025-10190-z

67. Richard V, Lorthe E, Dumont R, et al. Determinants and health-related consequences of screen time in children and adolescents: post-COVID-19 insights from a prospective cohort study. Swiss Med Wkly. 2025;155:4247. doi:10.57187/s.4247

68. Mattson G, Kuo DZ, COMMITTEE ON PSYCHOSOCIAL ASPECTS OF CHILD AND FAMILY HEALTH, et al. Psychosocial Factors in Children and Youth With Special Health Care Needs and Their Families. Pediatrics. 2019;143(1):e20183171. doi:10.1542/peds.2018-3171

69. Warschburger P, Petersen AC, von Rezori RE, et al. A prospective investigation of developmental trajectories of psychosocial adjustment in adolescents facing a chronic condition - study protocol of an observational, multi-center study. BMC Pediatr. 2021;21(1):404. doi:10.1186/s12887-021-02869-9

70. Pinquart M. Health-Related Quality of Life of Young People With and Without Chronic Conditions. Journal of Pediatric Psychology. 2020;45(7):780–792. doi:10.1093/jpepsy/jsaa052

71. Pinquart M, Shen Y. Depressive Symptoms in Children and Adolescents with Chronic Physical Illness: An Updated Meta-Analysis. Journal of Pediatric Psychology. 2011;36(4):375–384. doi:10.1093/jpepsy/jsq104

72. Lorthe E, Dumont R, Richard V, et al. Well-Being of Children and Adolescents with and without Special Health Care Needs Following the Lifting of Pandemic-Related Restrictions. J Pediatr. 2025;281:114528. doi:10.1016/j.jpeds.2025.114528

73. Richard V, Lorthe E, Dumont R, et al. Mental health trajectories of children and adolescents up to five years after the onset of the COVID-19 pandemic. medRxiv 2025.11.30.25340982; doi: 10.64898/2025.11.30.25340982.

74. Richard V, Lorthe E, Dumont R, et al. Psychosocial factors mediate social inequalities in health-related quality of life among children and adolescents. BMC Public Health. 2024;24(1):2986. doi:10.1186/s12889-024-20393-0

75. Bovio N, Richard V, Lorthe E, et al. Health behaviors, family context and school difficulties as mediators of social inequalities in school-related well-being and anxiety. In preparation.

76. Dumont R, Blanchard-Rohner G, Richard V, et al. The interplay of social and family environments in shaping adolescent health behaviours: a population-based study. Public Health. 2025;248:105931. doi:10.1016/j.puhe.2025.105931

77. Lorthe E, Richard V, Dumont R, et al. Socioeconomic conditions and children’s mental health and quality of life during the COVID-19 pandemic: An intersectional analysis. SSM Popul Health. 2023;23:101472. doi:10.1016/j.ssmph.2023.101472

78. Galea S, Tracy M. Participation Rates in Epidemiologic Studies. Annals of Epidemiology. 2007;17(9):643–653. doi:10.1016/j.annepidem.2007.03.013

